# Trilingual (EN/ZH-CN/JP) synthetic dataset of cerebral infarction patient-nurse bedside dialogs with metadata

**DOI:** 10.64898/2026.01.05.25342252

**Authors:** Kazuya Taira, Shuntaro Yada, Takahiro Itaya, Soshiro Ogata, Eri Kiyoshige, Ayame Hanada

## Abstract

We propose a large-scale synthetic dataset that correlates structured background information aligned with the actual distribution of patients with cerebral infarction, nurse characteristics, and nurse-patient dialogues across diverse scenarios. Medical dialogue corpora are scarce due to privacy and access restrictions. Even when available, they primarily focus on physician-patient interactions and offer limited metadata (clinical covariates, staff characteristics, etc.). To address this gap, this resource conditions large language models on patient covariate tables and nurse characteristics, generating multilingual, daily-structured dialogues (7 scenarios per day) using a standardized JSONL schema. Potential applications include nursing education under conditions of limited clinical practice time (supporting scenario-based therapeutic communication and formative assessment), research involving controlled experiments on nursing record formats and quality indicators, and training or benchmarking AI models when actual texts cannot be shared (subject to the selected model’s terms of use). All content is synthetic data and does not contain protected health information. Reproducible scripts pair patients with nurses, assign care pathways, and generate conversations on a scale.

## Introduction

We constructed a large-scale demo dataset that includes the background of patients with cerebral infarction, nurse attributes, and nurse-patients conversation dialogues. The training resources that integrate structured patient background, medical staff attributes, and realistic patient-medical staff conversations remain challenging. In the healthcare field, there are a few public dialogue corpora due to privacy and access constraints, and the few existing dataset tend to focus on medical staff-patient dialogue with limited meta-data.^1-2^ For instance, He, X. et al. reported that large-scale physician-patient dialogues, called “MedDialog” in English and Chinses, but the dataset has not linked patients’ clinical covariates and not included the detailed traits of physicians^3^. Additionally, the situation with dialogue data between nurses and patients is more challenging. ^1-2^ To solve these problems, synthetic generation with large language models (LLMs) offers a practical route to produce multilingual conversational data conditioned on structured tabular context, filling gaps where real data are unavailable or cannot be shared. ^1-3^

This dataset could have potential applications, such as educational setting for nurse training, future research on nursing documentation methods, and in training Artificial Intelligence (AI), including generative AI. In undergraduate nursing education, faculty shortages, limited clinical hours/placements, and patient-rights and privacy constraints contribute to reduced patient contact and educational barriers. ^4^ A large-scale randomized study by the National Council of State Boards of Nursing demonstrated that replacing part of clinical time with simulation can achieve equivalent educational outcomes, emphasizing the importance of high-quality, scenario-based learning materials when direct clinical opportunities are limited.^5^ A corpus including patient background variables, nurse characteristics, and multi-scenario daily nurse-patient conversation dialogues holds potential educational value for clinical practice records, nursing care communication exercises, and formative assessment.^4-5^

In addition, nursing documentation itself remains diverse. The previous review articles highlight ongoing efforts to standardize through recognized nursing terminologies and structured electronic records, while noting variations in completeness, structure, and adherence to recommended practices across different environments.^6-8^ Synthetically generated datasets with conversations annotated against patient covariates and care processes can support controlled experiments on documentation formats, content frameworks, and quality metrics, as research continues to evolve. ^6-8^

Finally, the use of generative AI to create clinical records and conversations is rapidly expanding. Several studies have shown that synthetic clinical text can be shared in data-scarce situations and is useful for training task-specific clinical models.^9^ At the same time, users must review the terms of use of model providers before reusing generated content to train other models. For example, OpenAI’s terms of service prohibit “developing models that compete with OpenAI”.^10^

## Methods

This data was generated through five processes: 1) Generating Japanese cerebral infarction patient background data, 2) Generating nurse characteristic data, 3) Generating patient treatment outcome data, 4) Generating patient-nurse pairs from the generated data, and 5) Generating conversation dialogues between patients and nurses within the pairs. This study exclusively used fully synthetic data generated to reproduce epidemiological statistical distributions and did not involve any real individuals or personally identifiable information. As no human subjects data were collected or analyzed, ethical approval and informed consent were not required.

### 1) Generating Japanese cerebral infarction patient background data (01_patient_bg.py)

Patient background data was adjusted to align with the target peripheral distribution based on published statistics in Japan Stroke Data Bank Report 2024^11^, generating a synthetic cohort with n=10,000. First, gender and age were generated to match the distribution shapes of target statistics. For males, a truncated normal distribution (median 75, IQR 15) was used for ages 18-99. For females, the median/IQR of a Beta distribution were numerically adjusted to match and linearly transformed to fit the age range. Next, for the NIHSS severity index, we constructed a logit linear predictor from age and gender. We adjusted the threshold (τ_1_, τ_2_) via binary search to assign the entire dataset such that P(NIHSS≤5) and P(NIHSS≤9) matched predetermined ratios. The 0–5 point distribution for mild cases (≤5) was assigned using the maximum residual method, adjusted to satisfy Q1=1, Q2=3, Q3=9. Subsequently, pre-modified Rankin Scale(pre-mRS) care level, medical history, smoking status, JCS, medications at admission, in-hospital procedures/complications, and NIHSS progression were generated using logistic/proportional odds approximation with age, sex, and NIHSS as explanatory variables. The intercept or cut-off point was calibrated to match the target rate for each item.

### 2) Generating nurse features data (02_nurse_features.py)

The characteristics of nurses vary significantly by country, region, and facility. Therefore, this study predefined a hypothetical distribution for demonstration purposes to prevent the occurrence of extremely rare categories, and generated synthetic data according to this distribution. The subjects were registered nurses (RNs) working in general wards, categorized by educational background (Diploma/ADN 35%, BSN 55%, MSN+ 10%), shift patterns (day 45%, evening 17%, night 23%, rotating 15%), work style (task_focused 25%, patient_centered 35%, by_the_book 20%, adaptive 20%), and communication style (supportive 35%, directive 20%, informational 25%, motivational 20%) were assigned via category sampling. All participants were assigned BLS certification. Additionally, ACLS (30%), wound_care (25%), and oncology_cert (15%) were assigned independently.

Years of clinical experience were sampled from a log-normal distribution, clipped to 0–40 years, rounded, and then classified using threshold rules according to Benner’s proficiency levels (novice < 1 year, advanced_beginner < 2 years, competent < 5 years, proficient < 10 years, expert ≥ 10 years). Personality traits (Big Five: O, C, E, A, N) were generated from multivariate normal distributions based on assumed means, variances, and correlations, then clipped to the range 0–1. Furthermore, continuous interpersonal skill-related indicators (empathy, assertiveness, patience, teaching, and teach-back usage probability) were generated from normal distributions with assumed means and variances, then clipped to 0–1.

As a communication-related derived variable, speech rate was clipped to 110–200 words/minute by applying style-specific biases (supportive −10, directive +5, informational 0, motivational −5) to the baseline of 150±20 words/minute. Motivational Interviewing (MI) proficiency was calculated by mapping Big Five traits (particularly Openness, Agreeableness, Conscientiousness, etc.) weights and minor noise onto a 0–1 scale. Compatibility with communication styles was also evaluated on a 0–1 scale using style-specific weighting (e.g., supportive style emphasizes Agreeableness and moderate Extraversion; directive style emphasizes Conscientiousness and Extraversion). Burnout risk was assigned 1 point each for night/shift work and novice/advanced-beginner skill level, with a total score of 0 classified as low, 1 as medium, and 2 as high.

### 3) Generating treatment outcome variables(03_patient_outcomes.py)

JCS was flagged as 0/non-zero, with blood glucose ≥180 mg/dL considered hyperglycemia and Cr ≥1.2 mg/dL considered renal impairment. tPA/thrombus retrieval (MT), onset-to-needle time, and rehabilitation within 48 hours (early rehab) were used as binary or thresholded variables.

Next, we constructed the severity score as a baseline severity indicator. The severity score was calculated as a weighted linear combination of age, NIHSS, JCS flag, atrial fibrillation, hypertension, diabetes, prior stroke, hyperglycemia, renal impairment, and pre-mRS, with treatment-related protective factors (tPA, MT, onset-to-treatment <90 minutes, early rehabilitation) added with negative weights (higher values indicate worse prognosis). Based on this severity score, the probability of in-hospital complications (in-hospital death, pneumonia, sICH, DVT, UTI) was calculated using a logistic function, and occurrence was determined by Bernoulli sampling. Only for sICH, tPA/MT, hyperglycemia, and systolic blood pressure ≥180 mmHg were considered, with an upper limit set on the probability to prevent it from becoming abnormally high.

Length of Stay (LOS) was modeled using a negative binomial distribution with the expected value given by exp(linear predictor). The linear predictor included S and each complication (increasing direction), tPA/MT/early rehab (decreasing direction), with the minimum value rounded to 4 days. The discharge destination was modeled as a multivalued logit with options: home / rehab hospital / nursing facility / acute transfer. Log odds were calculated based on S, age, prolonged hospitalization (>21 days), and presence of pneumonia or sICH. One option was sampled from the normalized probabilities. If an in-hospital death occurred, the discharge destination was overwritten to “death”.

The discharge mRS score was created as a latent continuous score comprising S and complications (pneumonia, sICH, DVT, UTI), tPA/MT, early rehabilitation, and pre-mRS. After adding Gaussian noise, it was mapped to an ordered category of 0–6 at a predetermined threshold (cut-off point) (overwritten as 6 in case of death). The 30-day readmission probability was estimated using logistic regression with explanatory variables including age ≥80 years, any in-hospital complication, discharge destination not home, and prolonged hospitalization (>21 days), and assigned a 0/1 value by random assignment. The 90-day mRS was calculated by determining the degree of improvement relative to the discharge mRS (improvement due to tPA/MT/early rehab, deterioration due to the severity score or complications + noise). If the calculated value fell below or exceeded a predetermined threshold, it was assigned a −1 / +1 step change, respectively; otherwise, it was assigned 0 steps. This was then clipped to the range 0–6 (fixed at 6 in case of death).

### 4) Creation of a patient-nurse paired dataset(04_pair_patients_nurses.py)

The character of nurses were limited to registered nurses in general wards, and the number of patients they could care at the same time was determined by their proficiency level according to Benner’s model: novice:3, advanced_beginner:4, competent:5, proficient:6, expert:7. The care quality score oriented by nurse was defined as below. Standardized years of nursing experience were defined between 0-1 by capping experience years at 20 and dividing by 20, while Benner bonus points assign 1.0 for “proficient”/”expert” and 0.5 for all other levels.

> The care quality score = 0.35× MI proficiency + 0.25 × Communication style alignment + 0.20 × Standardized years of nursing experience+ 0.20 × Benner’s score

We assigned three latent parameters to each patient as features independent of actual data: ( i ) initial severity, ( ii )social barriers, and ( iii ) recovery potential. These were sampled as random numbers from beta or normal distributions and handled within a range of 0 to 1. This expressed the diversity of patients at the time of allocation.

For each patient, two staff members were selected: a primary and a secondary. Selection followed a simple load balancing approach: ( i ) staff were ranked in descending order based on their load ratio (current load divided by capacity), and ( ii ) if load ratios were equal, seniority was used as the tiebreaker. Staff were then selected from the top of this list. After assignment, the primary caregiver’s load was increased by two cases and the secondary caregiver’s load by one case. This ensured load distribution progressed for subsequent patient assignments.

Finaly, we created one row per patient ID containing the following: primary and secondary nurse IDs, each nurse’s years of experience, Benner level, and care quality score; the patient’s latent parameters; and (if available in the input) length of stay and outcome.

### 5) Generating nurse-patient dialogue data (05_generate_dialogues_EN.py/05_generate_dialogues_ZH-CN.py/05_generate_dialogues_JP.py)

In this study, we employed a script that automatically generates seven daily patient-nurse interactions per patient. Generation was performed by issuing daily instructions to a large language model (LLM), using as inputs: (i) a base prompt template, and (ii) a correspondence table (pair information) between the patient and their assigned nurse.

The generation prompt were composed of two messages: system and user. On the user side, seven daily scenarios (upon waking/after breakfast/before lunch/after lunch / before dinner / before bedtime / night patrol) were fixed. The first utterance in each scene (a nurse’s fixed phrase) was explicitly specified. Each scene was set to 10– 14 turns initiated by the nurse, with continuation of the previous scene’s context. Variable parameters are patient_id, day, primary_nurse_id, and the day’s focus and events based on the care arc. As a representative example, the actual message input to the LLM is provided as Textbox1 when patient_id=“P001”, day=3, primary_nurse_id=“N123”, focus=“stabilization and adjustment of off-bed/pain/anxiety”, events=None. However, for the last day of the output period, specifications were set to add special final-day instructions (such as safety guidance, listening to concerns, or calm conversations) depending on events like discharge, transfer, or end-of-life care.

Additionally, we created three prompt variations—Japanese, English, and Chinese—for input to the LLM and confirmed that the output generates dialogues in each language.

Textbox 1. The sample prompt for generating dialogue (English version)

[User]

You are given a patient–nurse daily dialog task. Generate **seven scenes in one shot** for the specified day.

**Output must be JSON only**. Top level schema:

{“day”: number, “scenes”: [ {“id”: string, “conversation”: [ {“role”:”Nurse”,”text”:”…”},

{“role”:”Patient”,”text”:”…”} … ]}, … ] }

**Conversation rules (apply to every scene):**

- 10–14 utterances per scene, **starting with Nurse**.
- The **first Nurse sentence must use the fixed opening text exactly as written** for that scene.
- Avoid mechanical repetition; continue the narrative from the previous scene.
- Language: English only for all natural language (JSON keys/IDs may be in English as-is).

**Task context (fixed for this request):**

- patient_id: P001
- day: 3
- primary_nurse_id: N123
- Today’s Focus: stabilization and adjustment of off-bed/pain/anxiety
- Today’s Events: None

**Seven scenes to generate (with assignment, time, and fixed opening):**

- wake (When waking up) — time=06:30, assign=primary Fixed Nurse: “Good morning. Did you sleep well? How are you feeling this morning?”
- post_breakfast (After breakfast) — time=08:45, assign=primary Fixed Nurse: “How are you feeling after breakfast? Did you have a good meal?”
- pre_lunch (Before lunch) — time=11:30, assign=primary Fixed Nurse: “I’m here to check in before lunch. How has your condition changed this morning?”
- post_lunch (After lunch) — time=13:30, assign=primary Fixed Nurse: “I’m checking in after lunch. Did you have a good meal?”
- pre_dinner (Before Dinner) — time=17:30, assign=primary Fixed Nurse: “This is a patrol before dinner. Have there been any changes in your condition this afternoon?”
- lights_out (Ready to sleep) — time=21:00, assign=secondary Fixed Nurse: “Good evening. I’m here to check on you before lights out. Is there any change?”
- night (Night patrol) — time=23:30, assign=secondary Fixed Nurse: “Excuse me, this is the night patrol. Are you sleeping well?”

**Reminder:** Output **JSON only** with the schema above; do not include explanations or Markdown.

As the data output method, scenes were extracted from the LLM’s responses and saved as JSON Lines (.jsonl) files, one scene per line, along with each scene’s ID, label, assigned category, time, and assigned nurse ID, organized per patient and per day. The design skips patients for whom responses had already been generated up to the final day. Additionally, to improve efficiency when generating large sample volumes, multiple patients were processed in parallel using a thread pool. An upper limit on requests per minute (RPM) was set to regulate API call intervals. The generative AI model name, parallelism level, RPM, number of samples, random seed, etc., can be specified via command-line arguments.

### Data Record

We deposited five primary records at [Trilingual (EN/ ZH-CN /JP) synthetic dataset of cerebral infarction patient–nurse bedside dialogs with metadata] (DOI: **10.5281/zenodo.17518507**). The overview of the dataset generation workflow was shown in Figure 1.

- patient_bg.csv — synthetic patient background (demographics, NIHSS, pre-mRS, etc.).
- nurse_features.csv — synthetic nurse attributes (background, Big Five, communication traits).
- patients_df.csv — synthetic in-hospital outcomes and discharge metrics.
- patient_nurse_pairs.csv — patient–nurse pairing with latent parameters and care-quality scores
- dialogues JSONL files (EN/ZH-CN/JP) — per patient 7-scene daily dialogues (patient_id, day, scene, conversation …).

**Figure 1.**
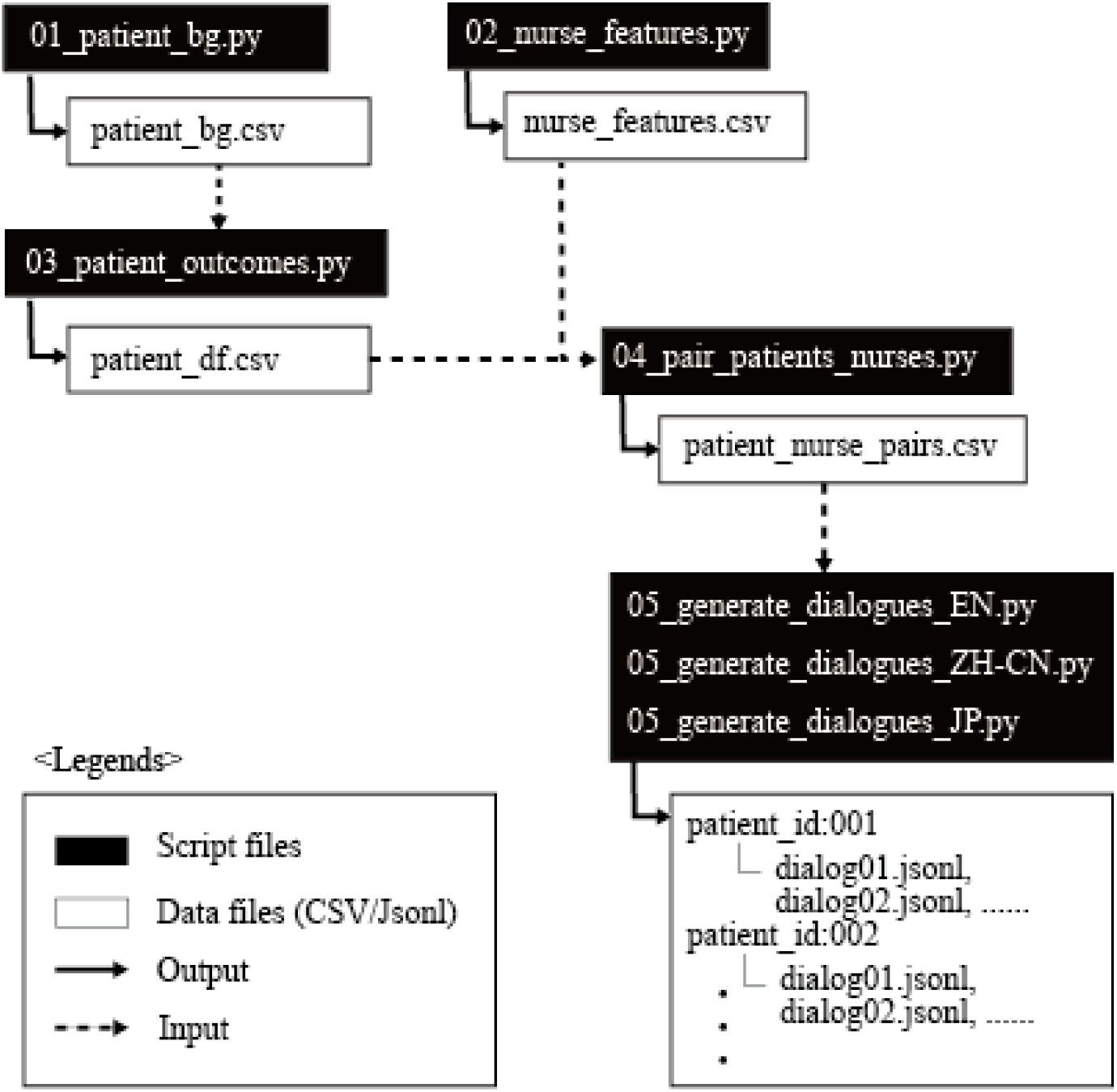
Overview of the dataset generation workflow

#### Formats and conventions

CSV files are UTF-8, comma-separated; missing values: NA (or empty). Times are local to Asia/Tokyo unless stated. Category vocabularies are enumerated in the data dictionary.

#### Data dictionary

A machine-readable codebook (codebook.csv) accompanies the deposit.

#### Versioning and reproducibility

Release v1.0 (2025-11-DD), generated with fixed random seeds (seed=42). Regeneration instructions are in README.md.

#### Access, license and citation

All records are available at DOI: **10.5281/zenodo.17518507** under MIT/CC4.0. Cite the dataset in the reference list and cite individual records in the Data Records section as numbered data citations.

### Technical Validation

All data are fully synthetic; no real patient or hospital information is included. We validated the release along three axes— distributional alignment/range integrity, cross-field logic, and reproducibility.

#### Distributional alignment & range integrity

The patient background variables were generated to match existing Japanese epidemiological data, confirming that the distributions and representative values were approximate (Table 1). The definitions of these variables were described in codebook.csv and we checked that the generated data and the codebook.csv were consistent with each other. Also, descriptive statistics such as mean, standard deviation, quartiles, maximum, and minimum values were calculated for all variables to check for any invalid values. Daily dialogue JSONL files were validated as a single valid JSON object per line containing required columns, ensuring no output in languages other than the specified ones (EN/ZH-CN/JP) and no mixing of multiple languages.

**Table 1.** The descriptive statistics of the generated variables and the target preexisted statistics.

|  | Target | Generated data |
| --- | --- | --- |
| Age, median [IQR] | 78 [70-85] | 77.7 [69.6-85.4] |
| Men, n (%) | 7,479 (57.3) | 5,750 (57.5) |
| NIHSS at admission, median [IQR] | 3 [1 - 9] | 3.5 [1.75 - 9.0] |
| Pre modified Rankin Scale, median [IQR] | 0 [0 - 2] | 0 [0-2] |
| Medical history, n (%) |  |  |
| Hearth failure | 4,130 (31.7) | 3170 (31.7) |
| Hypertension | 9,113 (69.9) | 6,950 (69.5) |
| Diabate | 3,468 (26.6) | 2720 (27.2) |
| Dyslipidemia | 5,089 (39.0) | 3908 (39.1) |
| Acute therapy, n (%) |  |  |
| Conservative therapy only | 10,581 (81.1) | 8142 (81.4) |
| Intravenous t-PA only | 842 (6.5) | 647 (6.5) |
| Endovascular treatment only | 969 (7.4) | 710 (7.1) |
| t-PA and endovascular treatment | 649 (5.0) | 501 (5.0) |

#### Cross logical check

We verified the consistency of rules between variables to the greatest extent possible. For example, if the discharge destination is death, modified Rankin Scale at discharge variable must be set to 6. Furthermore, since patient outcomes were generated based on NIHSS severity, we also confirmed that a consistent correlation was maintained between patient severity and treatment outcomes.

#### Reproducibility

In the variables generating process, We fixed random seeds(seed=42) to ensure reproductivity. Dialogue generation depends on the model API and is non-deterministic; to ensure reproducibility of results, the generated JSONL files are deposited alongside scripts.

#### Privacy & safety

The dataset is synthetic and contains no personally identifiable information. Identifiers are artificial and non-linkable.

## Data Availability

All data produced are available online at https://doi.org/10.5281/zenodo.17518507.

https://doi.org/10.5281/zenodo.17500646

## Data Availability

All data records are deposited at Repository: Trilingual (EN/ ZH-CN /JP) synthetic dataset of cerebral infarction patient–nurse bedside dialogs with metadata, DOI: 10.5281/zenodo.17518507. The deposit includes:

- patient_bg.csv (synthetic patient background),
- nurse_features.csv (synthetic nurse attributes),
- patient_df.csv (in-hospital outcomes and follow-up),
- patient_nurse_pairs.csv (patient–nurse pairings with latent parameters and care-quality scores),
- dialogs_run/…/*.jsonl (per-patient daily 7-scene dialogue files).

Variable definitions (types, units, vocabularies, provenance, derivations, and examples) are provided in codebook.csv. Unless otherwise noted, data are released under CC BY 4.0.

## Code Availability

All generation scripts are openly available at Zendo [doi: 10.5281/zenodo.17500646] under tag v1.0. The environment targets Python 3.10+ with primary dependencies: numpy, pandas, scipy, and openai (dialogue generation only). Minimal reproduction steps:

\# 1) Generate cerebral infarction patients character variables

python 01_patient_bg.py --n 10000 --seed 42 --out data/patient_bg.csv

\# 2) Generate nurse features variables

python 02_nurse_features.py --n-nurses 120 --seed 42 --output data/nurse_features.csv

\# 3) Generate patients treatment outcomes

python 03_patient_outcomes.py --input data/synth.csv --output data/patients_df.csv --seed 42

\# 4) Generate the patient–nurse pair data

python 04_pair_patients_nurses.py --patients data/patients_df.csv --nurses data/nurse_features.csv --output data/patient_nurse_pairs.csv --seed 42

\# 5) Generate dialogue data

$env:OPENAI_API_KEY = “xxxxx” ( Please replace xxxxx to your API key)

<English version>

python 05_generate_dialogues_EN.py --step1-path data/step1_messages_v5.json --index-csv data/patient_nurse_pairs_enriched.csv --out-dir data/dialogs_run --model gpt-5-mini --workers 8 --rpm 0 --sample-n 10 --seed 42

<Simplified Chinese version>

python 05_generate_dialogues_CN.py --step1-path data/step1_messages_v5.json --index-csv data/patient_nurse_pairs_enriched.csv --out-dir data/dialogs_run --model gpt-5-mini --workers 8 --rpm 0 --sample-n 10 --seed 42

<Japanese version>

python 05_generate_dialogues_JP.py --step1-path data/step1_messages_v5.json --index-csv data/patient_nurse_pairs_enriched.csv --out-dir data/dialogs_run --model gpt-5-mini --workers 8 --rpm 0 --sample-n 10 --seed 42

We provide requirements.txt for constructing the environment to support exact regeneration.

## Acknowledgement

This study was supported by the Japan Society for the Promotion of Science (JSPS KAKENHI 25K13935). The funder played no role in the study design, data collection, analysis, interpretation, or writing of the report.

## Author contributions

KT contributed to conceptualization, Methodology, Software, Funding Acquisition, Writing(lead), SY contributed to Software, and Supervision, TI contributed to Validation and Writing(support), SO contributed to Validation and Writing(support), EK contributed to Validation and Writing(support) and AH contributed to Validation and Writing(support).

## Competing Interests

The authors declare no competing interests.

## Notes

### Competing Interest Statement

The authors have declared no competing interest.

